# A cross-sectional study of socioeconomic status and treatment interruption among Japanese workers during the COVID-19 pandemic

**DOI:** 10.1101/2021.02.22.21252190

**Authors:** Kenji Fujimoto, Tomohiro Ishimaru, Seiichiro Tateishi, Tomohisa Nagata, Mayumi Tsuji, Hisashi Eguchi, Akira Ogami, Shinya Matsuda, Yoshihisa Fujino, for the CORoNaWork Project

**Author notes:** Address correspondence to Yoshihisa Fujino, M.D., M.P.H., Ph.D., Department of Environmental Epidemiology, Institute of Industrial Ecological Sciences, University of Occupational and Environmental Health, Japan, 1-1, Iseigaoka, Yahatanishiku, Kitakyushu, 807-8555, Japan. Author contributions: KF; Writing the manuscript, TI; creating the questionnaire, review of manuscripts, and advice on interpretation, TN; review of manuscripts and advice on interpretation, ST, HE, MT, KM and SM; Review of manuscripts, advice on interpretation, and funding for research, YF; Overall survey planning, creating the questionnaire, analysis, and drafting the manuscript.

## Abstract

**Background:** The COVID-19 pandemic has caused interruptions to chronic disease and non-emergency treatment. The purpose of this study is to examine which socioeconomic status groups are most at risk of treatment interruption.

**Methods:** This cross-sectional internet monitor study was conducted on December 22–26, 2020, when Japan experienced its third wave of COVID-19 infection. Out of a total of 33,302 participants in the survey, 9510 (5392 males and 4118 females) who responded that they required regular treatment or hospital visits were included in the analysis. A multilevel logistic model nested in the prefecture of residence was used to estimate the odds ratio (OR) for treatment disruption. We examined separate multivariate models for socioeconomic factors, health factors, and lifestyle factors.

**Results:** During a period of rapid COVID-19 infection, about 11% of Japanese workers who required regular treatment experienced interruptions to their treatment. The OR of treatment interruption associated with not being married compared to being married was 1.44; manual labor work compared to desk work was 1.30; loss of employment when the COVID-19 pandemic started and continued unemployment compared to being employed over the entire pandemic period was 1.62 and 2.57, respectively; and feeling financially unstable was 2.92.

**Conclusion:** Treatment interruption is a new health inequality brought about by COVID-19 with possible medium- and long-term effects, including excess mortality, morbidity, and productivity loss due to increased presenteeism. Efforts are needed to reduce treatment interruptions among workers who require regular treatment.

## Introduction

Soon after COVID-19 was first confirmed in China, it quickly spread around the world. In January 2020, health authorities confirmed that the infection had spread to Japan. Subsequently, the country experienced peaks of infection in May and August 2020, which lasted for relatively short periods of time. The number of infected people and deaths were considered low compared to that in other countries. However, in November 2020, the number of infections rose again, and in December, Japan was experiencing its third wave of infection. This time, a larger number of people were becoming infected compared to the previous two waves. The hospital bed occupancy rate was high, and medical treatment was being affected. In January 2021, the Japanese government declared a second state of emergency for 11 prefectures.

The COVID-19 pandemic has caused interruptions to chronic disease and non-emergency treatment around the world^1–5^. In the United States, 40% of adults are reportedly avoiding medical care^1^. According to a survey of 47 countries, only 14% of responding health care providers continued to provide normal face-to-face care^2^. In Japan, the number of prescriptions in May 2020 made up just 75% of the prescriptions given out one year prior in May 2019 ^3^. In pediatrics and otolaryngology, the number of prescriptions halved ^3^.

There are several reasons why treatment may be being interrupted during the COVID-19 pandemic. Because hospitals are perceived as high-risk places for infection, patients may be voluntarily abstaining from treatment because they are concerned about becoming infected in the hospital^1^. In addition, many medical institutions are rescheduling visits with non-emergency patients to enable them to deal with COVID-19 and to set up fever outpatient clinics^6,7^. It is also likely that the financial impact of COVID-19 is causing patients to discontinue treatment, given that financial difficulties generally lead to interruption of treatment for chronic diseases^8–10^.

Delay or avoidance of medical care may increase excess mortality directly or indirectly related to COVID-19^1^. Delays and avoidance of treatment can lead to worse management of chronic diseases, missed opportunities for regular checkups, and missed or delayed initiation of treatment for deteriorating health conditions. This results in an increase of morbidity and mortality from health conditions that are otherwise treatable or preventable^4,11,12^.

Furthermore, interruptions to treatment among workers increase presenteeism ^13^, a practice in which individuals continue to work while unwell. Continued presenteeism can lead to lower productivity, further deterioration of health status, and even difficulties with staying in the labor market^14^. Anxieties related to possible infection after seeking medical care and the need to reschedule regular medical appointments due to the COVID-19 pandemic are causing interruptions to medical treatment for workers. In addition, economic insecurity and job insecurity are causing workers to feel that they need to make their presence known to management, which can discourage workers from actively taking sick leave.

While treatment interruptions are increasing in Japan due to the COVID-19 pandemic^3^, it is unclear which groups of individuals are most at risk. COVID-19 is having a significant impact on the socioeconomic status of workers, and such changes are expected to affect the consultation behavior of workers with chronic diseases. Therefore, the purpose of this study is to examine which socioeconomic status groups are most at risk of treatment interruption.

## Methods

### Study Design and Subjects

This cross-sectional internet monitor study was conducted on December 22–26, 2020, when the third wave of COVID-19 infection started in Japan. Details of the protocol of this survey have already been reported^15^. Briefly, data were collected from workers who had employment contracts at the time of the survey, and were assigned by prefecture, job type, and sex. Out of a total of 33,302 people who participated in the survey, 27,036 people were included in the study after removing those found to have provided fraudulent responses. Of these, 9510 (5392 males and 4118 females) who responded that they required regular treatment or hospital visits were included in the present analysis.

### Assessment of treatment status

We used the following single-item question to examine the participants’ treatment status: “Do you have any disease that requires regular visits to the hospital or treatment?” Respondents were asked to select from the following options: “I do not have such a disease;” “I am continuing with hospital visits and treatment as scheduled;” “I am not able to continue with hospital visits and treatment as scheduled.”

### Assessment of socioeconomic status, health status, and lifestyle and work-related factors

The subjects provided responses to the questionnaire via the Internet. Socioeconomic factors included age, sex, marital status (married, unmarried, bereaved/divorced), occupation (mainly desk work, jobs mainly involving interpersonal communication, mainly labor), educational background, equivalent income (household income divided by the square root of household size), job change or unemployment after April 2020 (did not resign or change jobs; transferred to another company; resigned and entered into a new job immediately; unemployed for a period, but currently working; retired and started a business), and perception of financial situation (very difficult, slightly difficult, fairly difficult, comfortable).

Health and psychological factors included self-rated health, psychological distress, feeling alone, having a friend who can provide support, and having a health condition that requires company support to enable work. Psychological distress was assessed using Kessler 6 (K6)^16^, the validity of the Japanese version of which has been confirmed^17^. In the present study, a K6 score of 5 or higher was used as the cutoff for mild psychological distress. The following question was used to examine loneliness: “Have you ever felt alone?” Subjects chose from the following options: “never,” “a little,” “sometimes,” “usually,” “always.” Subjects were asked the following questions to determine if they had a health condition that required company support to enable work: “Do you require consideration or support from your company to continue working in your current health condition?” Subjects chose from the following three options: “no;” “yes, but I have not received support;” “yes, and I have received support.”

The following lifestyle and work-related factors were examined: smoking (never; quit smoking more than one year ago; quit smoking within the past year; started smoking less than one year ago; smoking for more than one year), alcohol consumption (6–7 days a week; 4–5 days a week; 2–3 days a week; less than 1 day a week; hardly ever), exercise habit, breakfast routine, time spent on one-way commute and overtime work hours per day. For exercise habit, subjects were asked to indicate the number of days per week for which they exercised for 30 minutes or more. For breakfast routine, subjects indicated the number days per week for which they ate breakfast.

### Statistical analysis

Age-sex adjusted odds ratios (ORs) and multivariate adjusted ORs were estimated using a multilevel logistic model nested in the prefecture of residence. We conducted separate multivariate analyses of socioeconomic factors, health factors, and lifestyle factors. For socioeconomic factors, the multivariate model was adjusted for sex, age, marital status, job type, equivalent household income, educational background, employment status, and financial comfort. For health-related factors, the multivariate model was adjusted for age, sex, self-rated health, psychological distress, feeling alone, presence of a friend who can provide support, and presence of a health condition that requires company support to enable work. For lifestyle-related factors and work-related factors, the multivariate model was adjusted for age, sex, smoking, alcohol consumption, exercise habit, breakfast routine, time spent on one-way commute and overtime work hours per day. All analyses used the incidence rate of COVID-19 by prefecture as a prefecture-level variable.

A *p* value less than .05 was considered statistically significant. All analyses were conducted using Stata (Stata Statistical Software: Release 16; StataCorp LLC, TX, USA).

## Results

Table 1 shows the basic characteristics of the survey subjects. Of the 9510 subjects who required regular treatment, 11% discontinued treatment. Those who were continuing treatment reported better self-rated health than those who had discontinued treatment. Those who had experienced treatment interruption were more likely to experience psychological distress (77%), feel alone (30.8%), and skip breakfast. Those who had interrupted treatment also worked more overtime hours.

**Table 1.**
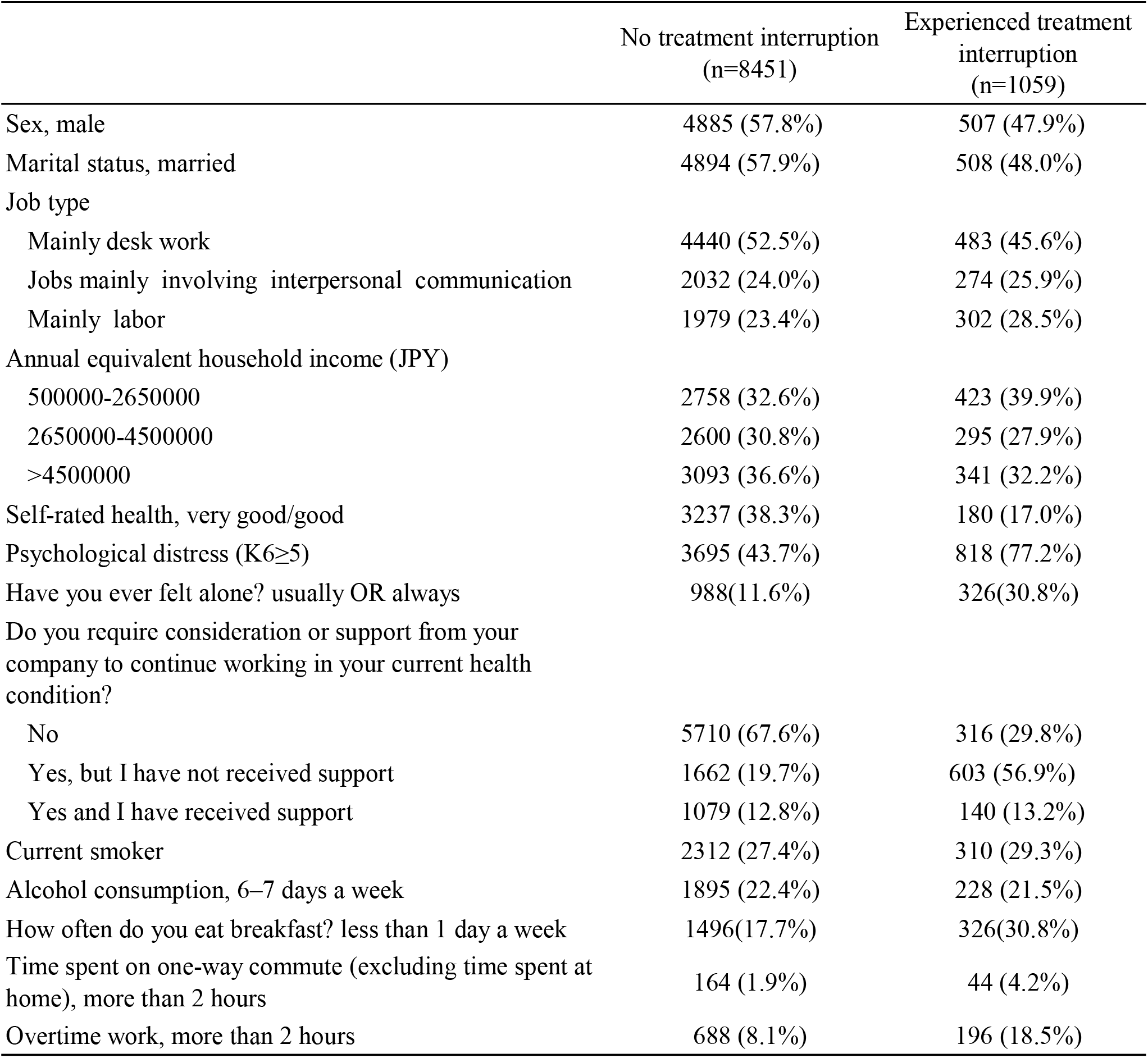
Basic characteristics of the study subjects

|  | No treatment interruption<br>(n=8451) | Experienced treatment<br>interruption<br>(n=1059) |
| --- | --- | --- |
| Sex, male | 4885 (57.8%) | 507 (47.9%) |
| Marital status, married | 4894 (57.9%) | 508 (48.0%) |
| Job type |  |  |
| Mainly desk work | 4440 (52.5%) | 483 (45.6%) |
| Jobs mainly involving interpersonal communication | 2032 (24.0%) | 274 (25.9%) |
| Mainly labor | 1979 (23.4%) | 302 (28.5%) |
| Annual equivalent household income (JPY) |  |  |
| 500000-2650000 | 2758 (32.6%) | 423 (39.9%) |
| 2650000-4500000 | 2600 (30.8%) | 295 (27.9%) |
| >4500000 | 3093 (36.6%) | 341 (32.2%) |
| Self-rated health, very good/good | 3237 (38.3%) | 180 (17.0%) |
| Psychological distress (K6≥5) | 3695 (43.7%) | 818 (77.2%) |
| Have you ever felt alone? usually OR always | 988(11.6%) | 326(30.8%) |
| Do you require consideration or support from your<br>company to continue working in your current health<br>condition? |  |  |
| No | 5710 (67.6%) | 316 (29.8%) |
| Yes, but I have not received support | 1662 (19.7%) | 603 (56.9%) |
| Yes and I have received support | 1079 (12.8%) | 140 (13.2%) |
| Current smoker | 2312 (27.4%) | 310 (29.3%) |
| Alcohol consumption, 6–7 days a week | 1895 (22.4%) | 228 (21.5%) |
| How often do you eat breakfast? less than 1 day a week | 1496(17.7%) | 326(30.8%) |
| Time spent on one-way commute (excluding time spent at<br>home), more than 2 hours | 164 (1.9%) | 44 (4.2%) |
| Overtime work, more than 2 hours | 688 (8.1%) | 196 (18.5%) |

Table 2 shows the association between socioeconomic status and treatment interruption. Marital status, job type, equivalent household income, employment status, and financial well-being were all associated with treatment interruption. Multivariate analysis showed that the OR of treatment discontinuation associated with not being married compared to being married was 1.44; manual labor compared to desk work was 1.30; loss of employment when the COVID-19 pandemic started and continued unemployment compared to being employed over the entire pandemic period was 1.62 and 2.57, respectively; and feeling financially unstable was 2.92.

**Table 2.** Association between socioeconomic status and treatment interruption

|  | Age-sex adjusted |  |  |  | Multivariate* |  |  |  |
| --- | --- | --- | --- | --- | --- | --- | --- | --- |
|  | OR | 95%CI |  | p | OR | 95%CI |  | p |
| Marital status |  |  |  |  |  |  |  |  |
| Married | reference |  |  |  | reference |  |  |  |
| Unmarried | 1.56 | 1.28 | 1.91 | <0.001 | 1.44 | 1.17 | 1.76 | <0.001 |
| Divorced/bereavement | 1.06 | 0.91 | 1.23 | 0.489 | 0.99 | 0.85 | 1.16 | 0.947 |
| Job type |  |  |  |  |  |  |  |  |
| Mainly desk work | reference |  |  |  | reference |  |  |  |
| Jobs mainly involving interpersonal communication | 1.16 | 0.99 | 1.36 | 0.071 | 1.14 | 0.97 | 1.33 | 0.116 |
| Mainly labor | 1.36 | 1.16 | 1.59 | <0.001 | 1.30 | 1.11 | 1.52 | 0.001 |
| Annual equivalent household income(JPY) |  |  |  |  |  |  |  |  |
| 500000-2650000 | 1.29 | 1.11 | 1.51 | 0.001 | 1.19 | 1.01 | 1.40 | 0.036 |
| 2650000-4500000 | 0.97 | 0.83 | 1.15 | 0.765 | 0.95 | 0.80 | 1.12 | 0.517 |
| >4500000 | reference |  |  |  | reference |  |  |  |
| Education |  |  |  |  |  |  |  |  |
| Junior high school | 1.27 | 0.75 | 2.13 | 0.376 | 1.01 | 0.60 | 1.72 | 0.967 |
| High school | 1.11 | 0.96 | 1.29 | 0.161 | 1.02 | 0.88 | 1.19 | 0.786 |
| Vocational school/college, university, graduate school | reference |  |  |  | reference |  |  |  |
| Have you resigned or changed jobs since April 2020? |  |  |  |  |  |  |  |  |
| Did not resign or change jobs | reference |  |  |  | reference |  |  |  |
| Transferred to another company | 1.00 | 0.52 | 1.92 | 0.996 | 0.97 | 0.50 | 1.88 | 0.932 |
| Resigned and entered into a new job immediately | 1.21 | 0.87 | 1.69 | 0.261 | 1.14 | 0.81 | 1.59 | 0.448 |
| Unemployed for a period, but currently working | 1.74 | 1.22 | 2.48 | 0.002 | 1.62 | 1.13 | 2.31 | 0.008 |
| Retired and started a business (e.g., running a company, personal business, or self-employment) | 2.76 | 1.75 | 4.36 | <0.001 | 2.57 | 1.63 | 4.07 | <0.001 |
| How do you feel about your current financial situation? |  |  |  |  |  |  |  |  |
| Very difficult | 3.10 | 2.39 | 4.02 | <0.001 | 2.92 | 2.25 | 3.80 | <0.001 |
| Slightly difficult | 1.50 | 1.16 | 1.92 | 0.002 | 1.44 | 1.12 | 1.85 | 0.005 |
| Fairly difficult | 0.97 | 0.75 | 1.24 | 0.804 | 0.95 | 0.74 | 1.22 | 0.690 |
| Comfortable | reference |  |  |  | reference |  |  |  |

\* The multivariate model was adjusted for sex, age, marital status, job type, equivalent household income, educational background, employment status, and perception of financial situation.

Table 3 shows the association between health status, psychological distress and treatment interruption. Multivariate analysis showed that the OR of treatment interruption associated with poor self-rated health was 5.27; experiencing psychological distress was 3.78; always feeling alone was 5.45; having no friends who can provide support was 2.13; and requiring company consideration to enable work but not receiving it was 5.87.

**Table 3.** Association between health status, psychological distress and treatment interruption.

|  | Age-sex adjusted |  |  |  | Multivariate* |  |  |  |
| --- | --- | --- | --- | --- | --- | --- | --- | --- |
|  | OR | 95%CI |  | p | OR | 95%CI |  | p |
| Self-rated health |  |  |  |  |  |  |  |  |
| Very good/good | reference |  |  |  | reference |  |  |  |
| Neither good nor bad | 1.86 | 1.54 | 2.23 | <0.001 | 1.85 | 1.54 | 2.23 | <0.001 |
| Bad | 5.23 | 4.36 | 6.27 | <0.001 | 5.27 | 4.39 | 6.34 | <0.001 |
| Psychological distress (K6≥5) | 3.84 | 3.29 | 4.47 | <0.001 | 3.78 | 3.24 | 4.41 | <0.001 |
| Have you ever felt alone? |  |  |  |  |  |  |  |  |
| Never | reference |  |  |  | reference |  |  |  |
| A little | 1.86 | 1.55 | 2.24 | <0.001 | 1.86 | 1.54 | 2.24 | <0.001 |
| Sometimes | 2.76 | 2.29 | 3.33 | <0.001 | 2.75 | 2.27 | 3.32 | <0.001 |
| Usually | 3.69 | 2.95 | 4.63 | <0.001 | 3.71 | 2.95 | 4.66 | <0.001 |
| Always | 5.39 | 4.37 | 6.65 | <0.001 | 5.45 | 4.40 | 6.76 | <0.001 |
| Do you have friends or neighbors with whom you can easily engage in small talk or daily conversation? No | 2.12 | 1.86 | 2.42 | <0.001 | 2.13 | 1.86 | 2.43 | <0.001 |
| Do you have someone you can ask for help? No | 2.32 | 2.03 | 2.64 | <0.001 | 2.31 | 2.02 | 2.64 | <0.001 |
| Do you require consideration or support from your company to continue working in your current health condition? |  |  |  |  |  |  |  |  |
| No | reference |  |  |  | reference |  |  |  |
| Yes, but I have not received support | 6.01 | 5.18 | 6.97 | <0.001 | 5.87 | 5.06 | 6.82 | <0.001 |
| Yes and I have received support | 2.07 | 1.67 | 2.56 | <0.001 | 2.06 | 1.66 | 2.54 | <0.001 |

\* The multivariate model was adjusted for age, sex, self-rated health, psychological distress, feeling alone, presence of a friend who can provide support, and presence of a health condition that requires company support to enable work.

Table 4 shows the association between lifestyle, occupational factors and treatment interruption. Smoking, drinking habit, breakfast habit, commuting time, and overtime work hours were associated with treatment interruption. In contrast, there was no association between exercise habit and treatment interruption. Multivariate analysis showed that the OR of treatment discontinuation associated with current smoking was 1.23; habitual drinking was 1.32; rarely eating breakfast was 2.28; a one-way commute of more than 2 hours was 1.86; and working more than 2 hours of overtime in an average day was 3.17.

**Table 4.** Association between lifestyle, occupational factors and treatment interruption.

|  | Age-sex adjusted |  |  |  | Multivariate* |  |  |  |
| --- | --- | --- | --- | --- | --- | --- | --- | --- |
|  | OR | 95%CI |  | p | OR | 95%CI |  | p |
| Current smoker | 1.28 | 1.10 | 1.48 | 0.001 | 1.23 | 1.06 | 1.43 | 0.006 |
| Alcohol consumption |  |  |  |  |  |  |  |  |
| 6–7 days a week | 1.12 | 0.94 | 1.34 | 0.214 | 1.32 | 1.06 | 1.65 | 0.014 |
| 4–5 days a week | 0.81 | 0.62 | 1.06 | 0.124 | 0.96 | 0.71 | 1.29 | 0.784 |
| 2–3 days a week | 0.80 | 0.64 | 1.01 | 0.057 | 0.95 | 0.73 | 1.24 | 0.706 |
| Less than 1 day a week | 0.85 | 0.70 | 1.03 | 0.101 | 1.16 | 0.95 | 1.40 | 0.139 |
| Hardly ever | reference |  |  |  | reference |  |  |  |
| How often do you exercise for 30 minutes or more per day? |  |  |  |  |  |  |  |  |
| 6–7 days a week | reference |  |  |  | reference |  |  |  |
| 4–5 days a week | 0.92 | 0.62 | 1.37 | 0.677 | 0.93 | 0.63 | 1.39 | 0.738 |
| 2–3 days a week | 0.92 | 0.65 | 1.31 | 0.662 | 0.95 | 0.67 | 1.35 | 0.771 |
| Less than 1 day a week | 0.96 | 0.68 | 1.35 | 0.819 | 0.98 | 0.70 | 1.38 | 0.918 |
| Hardly ever | 1.13 | 0.83 | 1.52 | 0.440 | 1.13 | 0.83 | 1.53 | 0.432 |
| How often do you eat breakfast? |  |  |  |  |  |  |  |  |
| 6–7 days a week | reference |  |  |  | reference |  |  |  |
| 4–5 days a week | 1.66 | 1.34 | 2.05 | <0.001 | 1.66 | 1.34 | 2.05 | <0.001 |
| 2–3 days a week | 1.89 | 1.47 | 2.44 | <0.001 | 1.86 | 1.45 | 2.41 | <0.001 |
| Less than 1 day a week | 1.73 | 1.29 | 2.31 | <0.001 | 1.71 | 1.27 | 2.29 | <0.001 |
| Hardly ever | 2.35 | 1.99 | 2.76 | <0.001 | 2.28 | 1.94 | 2.69 | <0.001 |
| Time spent on one-way commute (excluding time spent at home) |  |  |  |  |  |  |  |  |
| More than 2 hours | 1.77 | 1.22 | 2.58 | 0.003 | 1.86 | 1.27 | 2.71 | 0.001 |
| More than 1 hour | 0.93 | 0.74 | 1.18 | 0.560 | 0.99 | 0.79 | 1.25 | 0.950 |
| More than 30 minutes | 0.82 | 0.67 | 1.01 | 0.057 | 0.86 | 0.70 | 1.05 | 0.143 |
| Less than 30 minutes | 0.76 | 0.62 | 0.92 | 0.004 | 0.76 | 0.63 | 0.92 | 0.006 |
| Almost none | reference |  |  |  | reference |  |  |  |
| Overtime work |  |  |  |  |  |  |  |  |
| More than 2 hours | 2.95 | 2.43 | 3.59 | <0.001 | 3.17 | 2.61 | 3.87 | <0.001 |
| More than 1 hour | 1.35 | 1.11 | 1.64 | 0.003 | 1.44 | 1.18 | 1.75 | <0.001 |
| More than 30 minutes | 1.40 | 1.16 | 1.70 | 0.001 | 1.47 | 1.21 | 1.78 | <0.001 |
| Less than 30 minutes | 1.18 | 0.95 | 1.46 | 0.141 | 1.21 | 0.98 | 1.51 | 0.080 |
| Almost none | reference |  |  |  | reference |  |  |  |

\* The multivariate model was adjusted for age, sex, smoking, alcohol consumption, exercise habit, breakfast routine, time spent on one-way commute and overtime work hours per day.

## Discussion

This study showed that during the COVID-19 pandemic, 11% of workers in Japan who required regular medical attention discontinued treatment. Workers with poorer socioeconomic status, health status, and unfavorable lifestyle and working conditions were more likely to have experienced treatment discontinuation.

It is well known that socioeconomic status is an important factor in an individual’s ability to access to healthcare. Further, the circumstances of disadvantaged persons are exacerbated in emergencies such as disasters^8,18^. In this study, we showed that during the COVID-19 pandemic, workers with more disadvantaged socioeconomic status were more likely to experience interruptions to the medical treatment. In particular, individuals with low income, who experienced unemployment, and had poor financial stability were more likely to experience treatment interruption. These results are consistent with reports on the impact of economic deprivation on treatment discontinuation due to COVID-19 in other countries^1^. While we were unable to determine causality due to the cross-sectional nature of this study, it is possible that there is causality in both directions in the relationship between socioeconomic status and treatment discontinuation. Economic deprivation can be a direct reason for treatment interruption^8,18^. Workers in precarious employment situations may not have the right to sick leave or may be hesitant to take sick leave^13,19^, and thus give up treatment. On the other hand, interruptions to treatment may lead to exacerbation of health conditions, leading to job loss and reduced income.

In the present study, we showed that workers with poorer health status were more likely to experience treatment interruption: those with poorer self-rated health, psychological distress, loneliness, and workplace support were more likely to experience treatment interruption. The causal relationship between health status and treatment interruption may also be bi-directional. Several studies have reported that a major reason for treatment interruption is anxiety related to becoming infected when visiting a hospital^1,2,4^. In the present study, greater psychological distress was associated with higher odds of discontinuing treatment, suggesting that anxiety about infection may influence treatment discontinuation. In addition, subjects who did not receive company support were more likely to discontinue treatment. This may be because company support includes adjustment of an individual’s work hours for treatment and encouragement from management for the worker to see a doctor. Loneliness may also be related to whether or not a work colleague is able to accompany the subject to the doctor. Alternatively, treatment interruption may lead to deterioration of health status. Treatment interruption can increase the incidence of serious myocardial infarction ^4^. It is also possible that treatment interruption may lead to deterioration in health status and self-rated health.

The present study also examined the association of unfavorable lifestyle and work-related factors with treatment interruption. Unfavorable lifestyle habits may be related to health literacy^20,21^, which may in turn affect adherence to regular medical visits and treatment^22^. It is also conceivable that the relationship between lifestyle and treatment interruption may be confounded by socioeconomic status. Our findings also suggest that increased commute times and overtime work may reduce workers’ free time and hinder them from seeking medical attention, which is consistent with the findings of a previous study^23,24^.

Treatment discontinuation is a health disparity caused by COVID-19 and is an emerging public health issue. This study shows the relationship between socioeconomic status and treatment discontinuation in terms of health disparities. Regardless of the causal relationship, individuals with more disadvantaged socioeconomic status and health status were more likely to experience treatment interruption. It is therefore paramount that we prepare for the possibility that COVID-19 may not only threaten as an acute infectious disease, but also by affecting the prognosis and complications of those with chronic and lifestyle-related diseases through the socioeconomic environment. Campaigns to encourage the public to refrain from interrupting treatment are important. Relevant medical organizations and the Ministry of Health, Labor and Welfare in Japan are already conducting such campaigns. In this study, we found that workers with unfavorable lifestyle habits were more likely to experience treatment interruption. Therefore, a strategy to ensure that outreach messages are delivered efficiently is needed, especially to populations with low health literacy. Telemedicine is a promising method by which to handle treatment interruptions arising due to infection-related anxiety^25^. Extending the length of prescriptions is also a good approach. However, the safety of these approaches must be monitored, given that there are now fewer opportunities to observe patients’ health status. Our findings also demonstrate the importance of proactive support by companies to facilitate the delivery of treatment to sick workers. Such support is expected to reduce presenteeism and improve labor productivity.

Some limitations of this study warrant mention. First, we did not identify the reasons for treatment discontinuation. There are many possible explanations for why workers may discontinue treatment, including anxiety about COVID-19, financial reasons, lack of understanding of their disease, and lack of time to see a doctor. Second, we did not identify what diseases the workers were attempting to manage. Workers’ reasons for treatment interruption and the resulting effects are likely to be very different in the case of chronic diseases such as hypertension and diabetes, compared to cancer. Third, we did not inquire about the timing of treatment interruption; thus, we were unable to clarify the relationship between the timing of treatment interruption and changes in socioeconomic status, including employment and income, and lifestyle.

In conclusion, during a period of rapid COVID-19 infection, about 11% of Japanese workers who required regular treatment experienced treatment interruption. Disadvantageous socioeconomic status, poor health, and unfavorable lifestyle habits were associated with treatment interruption. Thus, treatment interruption is a new health inequality brought about by COVID-19 with possible medium- and long-term effects, including excess mortality, morbidity, and productivity loss due to increased presenteeism. Efforts are needed to reduce treatment interruption among workers who require regular treatment.

## Data Availability

Data not available due to ethical restrictions

## Acknowledgements

This study was funded by a research grant from the University of Occupational and Environmental Health, Japan; a general incorporated foundation (Anshin Zaidan) for the development of educational materials on mental health measures for managers at small-sized enterprises; Health, Labour and Welfare Sciences Research Grants: Comprehensive Research for Women’s Healthcare (H30-josei-ippan-002) and Research for the establishment of an occupational health system in times of disaster (H30-roudou-ippan-007); and scholarship donations from Chugai Pharmaceutical Co., Ltd.

Present members of the Collaborative Online Research on the Novel-coronavirus and Work (CORoNaWork) Project are: Dr. Yoshihisa Fujino (current chairperson), Dr. Akira Ogami, Dr. Arisa Harada, Dr. Ayako Hino, Dr. Chimed-Ochir Odgerel, Dr. Hajime Ando, Dr. Hisashi Eguchi, Dr. Kazunori Ikegami, Dr. Keiji Muramatsu, Dr. Koji Mori, Dr. Kyoko Kitagawa, Dr. Masako Nagata, Dr. Mayumi Tsuji, Dr. Rie Tanaka, Dr. Ryutaro Matsugaki, Dr. Seiishiro Tateishi, Dr. Shinya Matsuda, Dr. Tomohiro Ishimaru, Dr. Tomohisa Nagata, Dr. Yosuke Mafune, and Ms. Ning Liu, in alphabetical order. All members are affiliated with the University of Occupational and Environmental Health, Japan.

